# Comparing Stroke Risk in Patients Treated with Selective Serotonin Reuptake Inhibitors (SSRIs) versus Non-SSRI Antidepressants: A retrospective cohort study and meta-analysis

**DOI:** 10.1101/2025.10.28.25339028

**Authors:** Paco Wong Chun Yeung, Qi Sun, Jessie Ching Yan Su, Francisco Tsz Tsun Lai

**Affiliations:** Department of Pharmacology and Pharmacy, Li Ka Shing Faculty of Medicine, The University of Hong Kong, Hong Kong SAR; Advanced Data Analytics for Medical Science (ADAMS) Limited, Hong Kong SAR; Department of Family Medicine and Primary Care, School of Clinical Medicine, Li Ka Shing Faculty of Medicine, The University of Hong Kong, Hong Kong SAR

## Abstract

**Aims:** Despite the presence of studies indicating a potential elevated risk of stroke associated with selective serotonin reuptake inhibitors (SSRIs), current evidence is inconclusive. This study aims to evaluate the stroke risk associated with SSRI use using non-SSRI antidepressants as comparator through retrospective cohort study and meta-analysis of observational studies.

**Methods:** We extracted data from a territory-wide public healthcare database in Hong Kong to conduct a retrospective cohort study of patients aged 18+ years who started on SSRI or non-SSRI antidepressants between January 2018 to April 2024. Poisson regression with robust variance estimation was conducted to estimate the incidence rate ratio of stroke in SSRI users using non-SSRI users as active comparator. We subsequently conducted a systematic review and meta-analysis based on the current cohort study and all existing published observational data. Quality of studies was assessed using the Newcastle-Ottawa Scale.

**Results:** 122,679 individuals were included in the cohort study, among which 55,279 were SSRI users. SSRI users had an adjusted HR of 0.95 (95% CI 0.77-1.20) for stroke compared to non-SSRI users, suggesting a non-significant lower risk of stroke. Findings were consistent across subgroups by stroke types (i.e. ischemic stroke and hemorrhagic stroke). The result of our cohort study was aggregated with 5 other observational studies, and a pooled estimates of RRs were extracted (RR 0.93, 95% CI 0.81-1.07).

**Conclusion:** Our findings suggested that compared with non-SSRI antidepressants, SSRIs are not associated with a higher risk of stroke based on all available observational data.

## INTRODUCTION

Selective serotonin reuptake inhibitors (SSRIs) are among the most prescribed classes of antidepressants in recent years due to their effectiveness and good tolerability [1]. However, the use of SSRI has also been suggested to be a risk factor for stroke, including ischemic stroke and hemorrhagic stroke [2, 3, 4, 5], while a few showing a decrease in stroke risk [6]. This potential correlation is likely related to a disruption in hemostasis induced by SSRIs [7], in which SSRIs are suggested to reduce the ability of platelets to reuptake serotonin from bloodstream, potentially reducing their capacity for platelet adhesion [7].

The inconclusive results observed across various studies maybe attributed to the inability of observational studies to properly identify depression and other mental health disorders as a significant confounder. In fact, the predominant focus of existing studies lies in assessing stroke risk associated with SSRI use when compared to non-users, disregarding the underlying reasons (such as depression) that led to an individual being prescribed an antidepressant. Yet, several studies have provided support for an increased risk of stroke in persons suffering from depression [8, 9] and therefore could constitute an indication bias which may distort the results.

A recently published case-control study has recommended the use of non-SSRI antidepressants as an active comparator in future studies to minimize the impact of indication bias [10]. This is because people using the comparator drugs are typically those with similar metal health problems, i.e., similar indications. Therefore, we conducted a retrospective cohort study with an alternative methodology to assess the risk of stroke associated with antidepressant use, specifically by comparing SSRIs with non-SSRI antidepressants. The cohort study, along with other observational studies taking similar approaches, was subsequently reviewed systematically and meta-analyzed.

## METHOD

### Cohort study

#### Data source

In this study, we adopted the longitudinal territory-wide electronic health record database maintained by the Hospital Authority (HA), which is the primary provider of public inpatient services and major provider of outpatient services in Hong Kong [11]. This database essentially covers crucial details such as patients’ demographic profiles, consultation dates (including hospital admission and discharge dates), medical diagnoses, procedures, medication dispensing records, and results of laboratory tests. Each medical record is linked to a distinct anonymized reference key assigned to every individual patient, serving as a unique identifier for data integration across various healthcare facilities. Diagnoses are coded based on the International Classification of Diseases, Ninth Revision, Clinical Modifications (ICD-9-CM), and medications are coded according to codes stated in British National Formulary (B.N.F.) Notably, this database has previously been used in several high-quality, population-based studies [12–14], and has demonstrated high coding accuracy [15].

The study protocol was approved by the Institutional Review Board of the University of Hong Kong/ Hospital Authority Hong Kong West Cluster (reference number: UW 20-130). Informed consent was not required for this study because patient information used for secondary data analyses was strictly anonymized.

#### Study design and population

We adopted a retrospective cohort design for this study. New antidepressant users identified as those who used antidepressants (B.N.F. code 4.3.1 – 4.3.4) for the first time between January 2018 and April 2024 were included. Index date was designed as the date of first recorded prescription of antidepressants. Patients who (i) had a prior record of using antidepressants before age 18; (ii) those who used antidepressants for only one day; (iii) those who diagnosed stroke occurring before or within 30 days of starting antidepressants; and (iv) those who has record errors (e.g. lack of demographic information) were excluded. To select patients who use SSRI or non-SSRI only, patients who concurrently use SSRI and non-SSRI as initial antidepressant therapy were also excluded. All patients were followed from the index date until the (i) diagnosis of stroke; (ii) other diseases caused death; (iii) change of prescription to another antidepressant; (iv) discontinuation of initial type antidepressant, or (v) end of data availability (i.e., April 30, 2024), whichever occurred the earliest.

#### Exposure

Antidepressant use was the main exposure in the analysis. It was defined as the use of any antidepressant medication and was further categorized according to B.N.F. coding into a SSRIs group (B.N.F. 4.3.3) and a non-SSRIs group (B.N.F. 4.3.1, 4.3.2, 4.3.4). Patients were classified according to the type of antidepressants used on the index date.

#### Outcome

Any stroke diagnosis (ICD-9-CM codes 430, 431, 434, detail in S1 table) was adopted as the main outcome of this study. Secondary outcomes include hemorrhagic stroke (ICD-9-CM code 430 and 431) and ischemic stroke (ICD-9-CM codes 434).

#### Statistical analysis

Doubly robust Cox model was used to estimate the adjusted hazard ratio (AHR) with 95% confidence intervals (CI) of stroke and its subtypes between SSRIs and non-SSRIs antidepressants users, with an offset term to account for varying follow-up times for patients. The overlapping propensity score, based on inverse probability of treatment weighting (IPTW), was used to balance the characteristics between the SSRI and non-SSRI users. The score was estimated by logistic regression, considering covariates including age, baseline comorbidities (major depressive disorder, hypertension, alcohol abuse, diabetes, sepsis, hyperlipidemia, heart failure, atrial fibrillation, lung cancer, systemic lupus erythematosus, chronic obstructive pulmonary disease, rheumatoid arthritis, anxiety disorders, chronic kidney disease, pancreatic cancer, peripheral artery disease, coronary artery disease, obesity, amphetamine abuse, cocaine abuse, gastrointestinal cancers, tobacco use disorder, brain tumors, hematologic malignancies, carotid artery stenosis, migraine with aura, head and neck cancers), and prior use of medications (nonsteroidal anti-inflammatory drugs, statins, ACE inhibitors, Calcium channel blockers, Metformin, Atypical Antipsychotics, Combined hormonal contraceptives, Antiplatelet Agents, ARBs (Angiotensin-II receptor antagonists), Beta-adrenoceptor blocking drugs, GLP-1 receptor agonists, Varenicline, Anticoagulants, Epoetin Alfa, Estrogen-containing HRT, Pseudoephedrine), which will be classified into 3 categories, namely cumulative exposure less than one year, one to four years, and over five years. **Appendix 1** shows the ICD-9-CM codes and B.N.F. codes to ascertain the covariates. The standardized mean difference (SMD) was calculated to identify potential imbalances between SSRIs and non-SSRIs antidepressants users, with SMD greater than 0.1 indicating potential imbalance. Covariables showing imbalance were included in multivariable model for further adjustment. All data cleaning and analyses were conducted in R 4.4.0.

### Systematic review

#### Search Strategy and eligibility

The Preferred Reporting Items for Systematic Reviews and Meta-Analyses (PRISMA) statement is followed in the review [16]. A systematic search of articles published in English in scholarly journals was conducted in accordance with the protocol registered with PROSPERO (reference: CRD42024568935). 4 electronic databases, namely Cochrane Library, EMBASE, PubMed, and APA PsycINFO were used to perform the systematic search. The following search strategy structure was used: [SSRI] AND [Non-SSRI] AND [stroke] AND [Type of study]. The search strategy was executed on 9 August 2024. Search terms were derived from a systematic review published earlier [17]. Please refer to **Appendix 2** for details of search keywords and strategies.

This analysis included all published case-control and cohort studies investigating the association between SSRI and stroke risk from 2014 to 2024. The inclusion period is due to limitation noted in early observational studies, specifically the susceptibility to bias due to insufficient case representation or cohort size in exposure analyses [18–20]. Studies not published in English were excluded. Additionally, studies that (i) did not use non-SSRI as an active comparator, for instance, comparing stroke risk between SSRI users with non-users, (ii) had a study design that was neither cohort nor case-control, (iii) included subjects with any stroke history, or (iv) included non-human subjects were also excluded. To minimize selection bias, the research eligibility screening of the retrieved studies was independently carried out by two investigators, with discrepancies reconciled through discussion and consultation with senior author. For details of study eligibility, refer to **Appendix 3**.

#### Extraction

Screening of titles, abstracts, and full texts was conducted to evaluate if the studies identified during database search met specified inclusion criteria. Endnote Version 21 was used to store all retrieved studies and remove duplicate studies.

#### Quality assessment

Newcastle-Ottawa Scale (NOS) quality assessment tool was used to evaluate the quality of each included studies. The quality of studies was indicated by the number of stars, with nine stars indicating highest methodological quality. Please refer to **Appendix 4** for details of quality assessment.

#### Pooled estimates

Meta-analysis of the estimates of the association estimates and risk ratios (RR) were conducted after a satisfactory assessment result with regards to multivariable adjustment according to NOS. The estimates of the association between SSRI or non-SSRI and stroke risk were pooled using random-effect model. Exposure was binary-operationalized, comparing SSRI use to non-SSRI use. I^2^ statistic was used to analyze the fraction of variability that was due to heterogeneity. Cochrane Collaboration Review Manager (version 5.4.1) was used to generate pooled estimates and check for heterogeneity.

## RESULTS

### Cohort study

As shown in **Fig. 1**, between January 1, 2018, to April 30, 2024, a total of 141,953 people who started antidepressant use were identified from the database. Following exclusion of ineligible participants, the cohort size was 122,679, in which 55,279 were SSRI users and 67,400 were non-SSRI users.

**Fig. 1.**
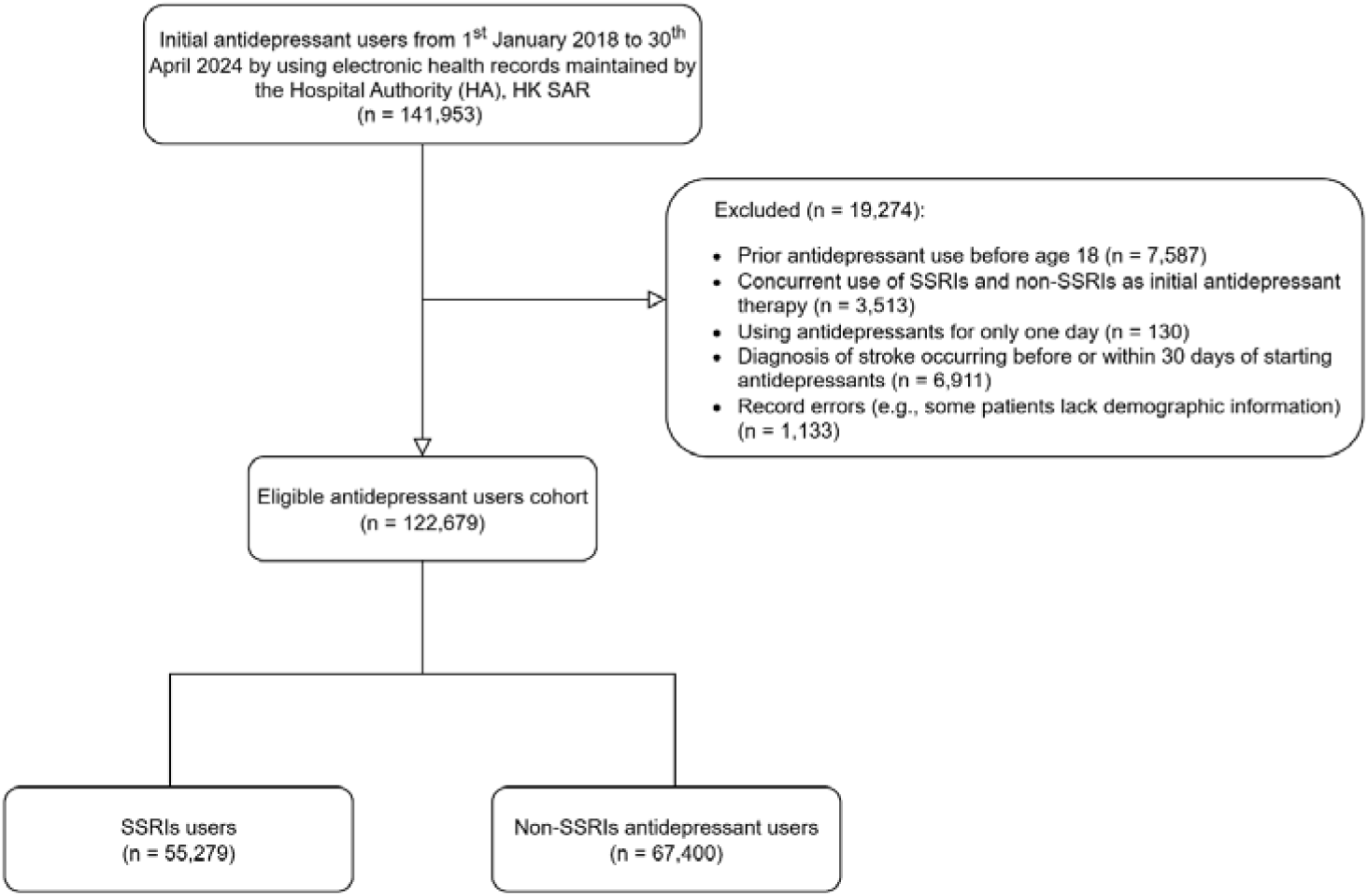
Flow chart of cohort selection process

### Cohort characteristics

**Table 1** shows the baseline characteristics of the cohort before and after applying IPTW. Before weighting, the mean (SD) age of SSRI group was 53 (20) years, which was approximately 10 years younger than that of non-SSRI group. The mean (SD) follow-up was 349 (393) days for SSRI group and 263 (356) days for non-SSRI group. The proportion of male patients taking SSRI was 37.7% while that of non-SSRI group was 32.0%. Notable differences were observed in types of baseline comorbidities. Specifically, hypertension (8.1% versus 13.1%) and diabetes (4.6% versus 7.8%) were more common comorbidities among non-SSRI users than SSRI users at baseline. After weighting, most of the baseline characteristics had SMD of less than 0.1 except age, sex, and some baseline comorbidities such as hypertension, diabetes, sepsis and lung cancer. These characteristics were adjusted in subsequent analyses to ensure well-balanced weighted cohort.

**Table 1.** Baseline characteristicsa.

|  | Unweighted |  |  | Weighted by inverse probability of treatment |  |  |
| --- | --- | --- | --- | --- | --- | --- |
|  | Non-SSRI | SSRI | SMD<sup>b</sup> | Non-SSRI | SSRI | SMD<sup>b</sup> |
| N | 67,400 | 55,279 |  | 122,301.4 | 123305 |  |
| Age (mean, standard deviation) | 63.25<br>(18.37) | 53.02<br>(19.95) | 0.533 | 58.87<br>(19.330) | 58.96<br>(20.18) | 0.005 |
| Male | 25,393<br>(37.7) | 17,666<br>(32.0) | 0.12 | 43000.1<br>(35.2) | 43450.2<br>(35.2) | 0.002 |
| *Comorbidities at baseline* |  |  |  |  |  |  |
| Major Depressive Disorder | 507 (0.8) | 696 (1.3) | 0.051 | 1,206.1 (1.0) | 1,191.8 (1.0) | 0.002 |
| Hypertension | 8,816 (13.1) | 4,459 (8.1) | 0.164 | 13,300.9 (10.9) | 13,476.5 (10.9) | 0.002 |
| Alcohol Abuse | 73 (0.1) | 91 (0.2) | 0.015 | 161.7 (0.1) | 166.9 (0.1) | 0.001 |
| Diabetes | 5,283 (7.8) | 2,519 (4.6) | 0.136 | 7,829.8 (6.4) | 7,993.7 (6.5) | 0.003 |
| Sepsis | 2,301 (3.4) | 878 (1.6) | 0.117 | 3,207.2 (2.6) | 3,357.0 (2.7) | 0.006 |
| Hyperlipidemia | 3,031 (4.5) | 1,544 (2.8) | 0.091 | 4,598.9 (3.8) | 4,691.1 (3.8) | 0.002 |
| Heart Failure | 1,946 (2.9) | 822 (1.5) | 0.096 | 2,780.9 (2.3) | 2,835.0 (2.3) | 0.002 |
| Atrial Fibrillation | 2,334 (3.5) | 1,049 (1.9) | 0.097 | 3,391.0 (2.8) | 3,473.9 (2.8) | 0.003 |
| Lung cancer | 941 (1.4) | 234 (0.4) | 0.103 | 1,181.9 (1.0) | 1,338.4 (1.1) | 0.012 |
| SLE | 145 (0.2) | 75 (0.1) | 0.019 | 218.9 (0.2) | 215.6 (0.2) | 0.001 |
| COPD | 979 (1.5) | 347 (0.6) | 0.081 | 1,330.6 (1.1) | 1,371.5 (1.1) | 0.002 |
| RA | 269 (0.4) | 104 (0.2) | 0.039 | 376.1 (0.3) | 390.0 (0.3) | 0.002 |
| Anxiety Disorders | 283 (0.4) | 511 (0.9) | 0.062 | 793.3 (0.6) | 790.2 (0.6) | 0.001 |
| CKD | 1,344 (2.0) | 735 (1.3) | 0.052 | 2,105.4 (1.7) | 2,174.5 (1.8) | 0.003 |
| Pancreatic Cancer | 166 (0.2) | 28 (0.1) | 0.051 | 191.1 (0.2) | 217.7 (0.2) | 0.005 |
| Peripheral Artery Disease | 228 (0.3) | 72 (0.1) | 0.043 | 301.1 (0.2) | 309.1 (0.3) | 0.001 |
| Coronary Artery Disease | 397 (0.6) | 211 (0.4) | 0.03 | 611.6 (0.5) | 615.5 (0.5) | <0.001 |
| Obesity | 394 (0.6) | 277 (0.5) | 0.011 | 669.2 (0.5) | 681.6 (0.6) | 0.001 |
| Amphetamine Abuse | 138 (0.2) | 65 (0.1) | 0.022 | 203.6 (0.2) | 203.7 (0.2) | <0.001 |
| Cocaine Abuse | 27 (0.0) | 24 (0.0) | 0.002 | 54.9 (0.0) | 59.3 (0.0) | 0.001 |
| Gastrointestinal Cancers | 549 (0.8) | 186 (0.3) | 0.063 | 742.4 (0.6) | 798.6 (0.6) | 0.005 |
| Tobacco Use Disorder | 14 (0.0) | 5 (0.0) | 0.01 | 18.5 (0.0) | 15.7 (0.0) | 0.002 |
| Brain Tumors | 292 (0.4) | 111 (0.2) | 0.041 | 416.5 (0.3) | 501.5 (0.4) | 0.011 |
| Hematologic Malignancies | 258 (0.4) | 74 (0.1) | 0.049 | 333.8 (0.3) | 359.4 (0.3) | 0.003 |
| Carotid Artery Stenosis | 56 (0.1) | 18 (0.0) | 0.021 | 74.6 (0.1) | 77.0 (0.1) | 0.001 |
| Migraine with Aura | 4 (0.0) | 3 (0.0) | 0.001 | 6.9 (0.0) | 7.1 (0.0) | <0.001 |
| Head and Neck Cancers | 30 (0.0) | 5 (0.0) | 0.022 | 35.1 (0.0) | 36.1 (0.0) | <0.001 |
| *Prior use of medications*<br>*(classified according to*<br>*cumulative exposure)* |  |  |  |  |  |  |
| NSAIDs |  |  | 0.131 |  |  | 0.003 |
| < 1 year | 63,682 (94.5) | 53,680 (97.1) |  | 116,956.9 (95.6) | 117,843.3 (95.6) |  |
| 1-5 years | 3,560 (5.3) | 1,545 (2.8) |  | 5,131.8 (4.2) | 5,246.5 (4.3) |  |
| > 5 years | 158 (0.2) | 54 (0.1) |  | 212.7 (0.2) | 215.2 (0.2) |  |
| Statins |  |  | 0.224 |  |  | 0.001 |
| < 1 year | 48,821 (72.4) | 45,201 (81.8) |  | 93,561.7 (76.5) | 94,287.7 (76.5) |  |
| 1-5 years | 11,127 (16.5) | 6,252 (11.3) |  | 17,430.7 (14.3) | 17,584.6 (14.3) |  |
| > 5 years | 7,452 (11.1) | 3,826 (6.9) |  | 11,309.0 (9.2) | 11,432.8 (9.3) |  |
| ACE inhibitors |  |  | 0.16 |  |  | 0.003 |
| < 1 year | 60,118 (89.2) | 51,676 (93.6) |  | 111,472.2 (91.1) | 112,278.3 (91.1) |  |
| 1-5 years | 4,612 (6.8) | 2,261 (4.1) |  | 6,888.8 (5.6) | 6,994.2 (5.7) |  |
| > 5 years | 2,670 (4.0) | 1,251 (2.3) |  | 3,940.4 (3.2) | 4,032.4 (3.3) |  |
| Calcium channel blockers |  |  | 0.25 |  |  | 0.002 |
| < 1 year | 47,433 (70.4) | 44,771 (81.0) |  | 91,760.3 (75.0) | 92,400.5 (74.9) |  |
| 1-5 years | 12,572 (18.7) | 6,787 (12.3) |  | 19,386.8 (15.9) | 19,585.7 (15.9) |  |
| > 5 years | 7,395 (11.0) | 3,721 (6.7) |  | 11,154.4 (9.1) | 11,318.8 (9.2) |  |
| Metformin |  |  | 0.16 |  |  | 0.002 |
| < 1 year | 58,545 (86.9) | 50,742 (91.8) |  | 108,886.1 (89.0) | 109,712.1 (89.0) |  |
| 1-5 years | 5,465 (8.1) | 2,809 (5.1) |  | 8,283.1 (6.8) | 8,382.0 (6.8) |  |
| > 5 years | 3,390 (5.0) | 1,728 (3.1) |  | 5,132.2 (4.2) | 5,210.9 (4.2) |  |
| Atypical Antipsychotics |  |  | 0.021 |  |  | 0.002 |
| < 1 year | 64,661 (95.9) | 53,228 (96.3) |  | 117,491.8 (96.1) | 118,428.1 (96.0) |  |
| 1-5 years | 1,882 (2.8) | 1,362 (2.5) |  | 3,273.4 (2.7) | 3,336.9 (2.7) |  |
| > 5 years | 857 (1.3) | 689 (1.2) |  | 1,536.2 (1.3) | 1,540.0 (1.2) |  |
| CHC |  |  | 0.03 |  |  | 0.001 |
| < 1 year | 67,320 (99.9) | 55,145 (99.8) |  | 122,089.1 (99.8) | 123,089.3 (99.8) |  |
| 1-5 years | 73 (0.1) | 129 (0.2) |  | 200.3 (0.2) | 201.9 (0.2) |  |
| > 5 years | 7 (0.0) | 5 (0.0) |  | 12.0 (0.0) | 13.8 (0.0) |  |
| Antiplatelet Agents |  |  | 0.187 |  |  | 0.003 |
| < 1 year | 57,290 (85.0) | 50,325 (91.0) |  | 107,179.7 (87.6) | 107,972.7 (87.6) |  |
| 1-5 years | 6,414 (9.5) | 3,246 (5.9) |  | 9,706.1 (7.9) | 9,886.6 (8.0) |  |
| > 5 years | 3,696 (5.5) | 1,708 (3.1) |  | 5,415.6 (4.4) | 5,445.7 (4.4) |  |

**Table 1.** Baseline characteristics<sup>a</sup> (continued)
|  | Unweighted |  |  | Weighted by inverse probability of treatment |  |  |
| --- | --- | --- | --- | --- | --- | --- |
|  | Non-SSRI | SSRI | SMD<sup>b</sup> | Non-SSRI | SSRI | SMD<sup>b</sup> |
| ARBs |  |  | 0.121 |  |  | <0.001 |
| < 1 year | 60,402 (89.6) | 51,423 (93.0) |  | 111,411.9 (91.1) | 112,318.3 (91.1) |  |
| 1-5 years | 5,155 (7.6) | 2,852 (5.2) |  | 8,031.3 (6.6) | 8,108.9 (6.6) |  |
| > 5 years | 1,843 (2.7) | 1,004 (1.8) |  | 2,858.2 (2.3) | 2,877.8 (2.3) |  |
| Beta-blockers |  |  | 0.158 |  |  | 0.002 |
| < 1 year | 57,559 (85.4) | 50,025 (90.5) |  | 107,159.8 (87.6) | 107,967.8 (87.6) |  |
| 1-5 years | 6,353 (9.4) | 3,505 (6.3) |  | 9,890.2 (8.1) | 10,013.4 (8.1) |  |
| > 5 years | 3,488 (5.2) | 1,749 (3.2) |  | 5,251.4 (4.3) | 5,323.8 (4.3) |  |
| GLP-1 receptor agonists |  |  | 0.089 |  |  | 0.003 |
| < 1 year | 64,376 (95.5) | 53,721 (97.2) |  | 117695.8 (96.2) | 118600.8 (96.2) |  |
| 1-5 years | 2,380 (3.5) | 1,239 (2.2) |  | 3637.4 ( 3.0) | 3709.5 (3.0) |  |
| > 5 years | 644 (1.0) | 319 (0.6) |  | 968.2 ( 0.8) | 994.7 (0.8) |  |
| Varenicline |  |  | 0.002 |  |  | <0.001 |
| < 1 year | 67,398 (100.0) | 55,278 (100.0) |  | 2.9 (0.0) | 2.7 (0.0) |  |
| 1-5 years | 0 (0) | 0 (0) |  | 0 (0) | 0 (0) |  |
| > 5 years | 0 (0) | 0 (0) |  | 0 (0) | 0 (0) |  |
| Anticoagulants |  |  | 0.078 |  |  | 0.003 |
| < 1 year | 66,068 (98.0) | 54,712 (99.0) |  | 120,396.7 (98.4) | 121,357.2 (98.4) |  |
| 1-5 years | 1,073 (1.6) | 450 (0.8) |  | 1,529.2 (1.3) | 1,552.0 (1.3) |  |
| > 5 years | 259 (0.4) | 117 (0.2) |  | 375.5 (0.3) | 395.8 (0.3) |  |
| Epoetin Alfa |  |  | 0.016 |  |  | 0.002 |
| < 1 year | 67,032 (99.5) | 55,038 (99.6) |  | 121,680.6 (99.5) | 122,661.8 (99.5) |  |
| 1-5 years | 317 (0.5) | 208 (0.4) |  | 535.0 (0.4) | 554.8 (0.4) |  |
| > 5 years | 51 (0.1) | 33 (0.1) |  | 85.8 (0.1) | 88.3 (0.1) |  |
| Estrogen-containing HRT |  |  | 0.013 |  |  | <0.001 |
| < 1 year | 67,353 (99.9) | 55,219 (99.9) |  | 122,196.8 (99.9) | 123,199.3 (99.9) |  |
| 1-5 years | 38 (0.1) | 50 (0.1) |  | 87.3 (0.1) | 87.9 (0.1) |  |
| > 5 years | 9 (0.0) | 10 (0.0) |  | 17.3 (0.0) | 17.9 (0.0) |  |
| Pseudoephedrine |  |  | <0.001 |  |  | <0.001 |
| < 1 year | 67,400 (100.0) | 55,279 (100.0) |  | 122,301.4 (100.0) | 123,305.0 (100.0) |  |
| 1-5 years | 0 (0) | 0 (0) |  | 0 (0) | 0 (0) |  |
| > 5 years | 0 (0) | 0 (0) |  | 0 (0) | 0 (0) |  |

<sup>a</sup> Values are expressed as frequency (%) unless otherwise specified
<sup>b</sup> Standardized mean difference
SLE, Systemic Lupus Erythematosus; COPD, Chronic Obstructive Pulmonary Disease; RA, Rheumatoid Arthritis; CKD, Chronic Kidney Disease; NSAIDs, Non-Steroidal Anti-Inflammatory Drugs; ACE, Angiotensin-Converting Enzyme; CHC, Combined Hormonal Contraceptive; ARBs, Angiotensin II Receptor Blockers

#### Main analysis

Among all 122,679 patients exposed to SSRI or non-SSRI, a total of 434 patients were diagnosed with stroke. Of these cases, 154 were SSRI users (n = 55,279), while the remaining 280 were non-SSRI users (n = 67,400). **Fig. 2** illustrates the weighted cumulative incidence of stroke over the follow-up period for SSRI and non-SSRI users. Notably, the incidence of stroke among non-SSRI users consistently higher than that among SSRI users throughout the entire follow-up period.

**Fig. 2.**
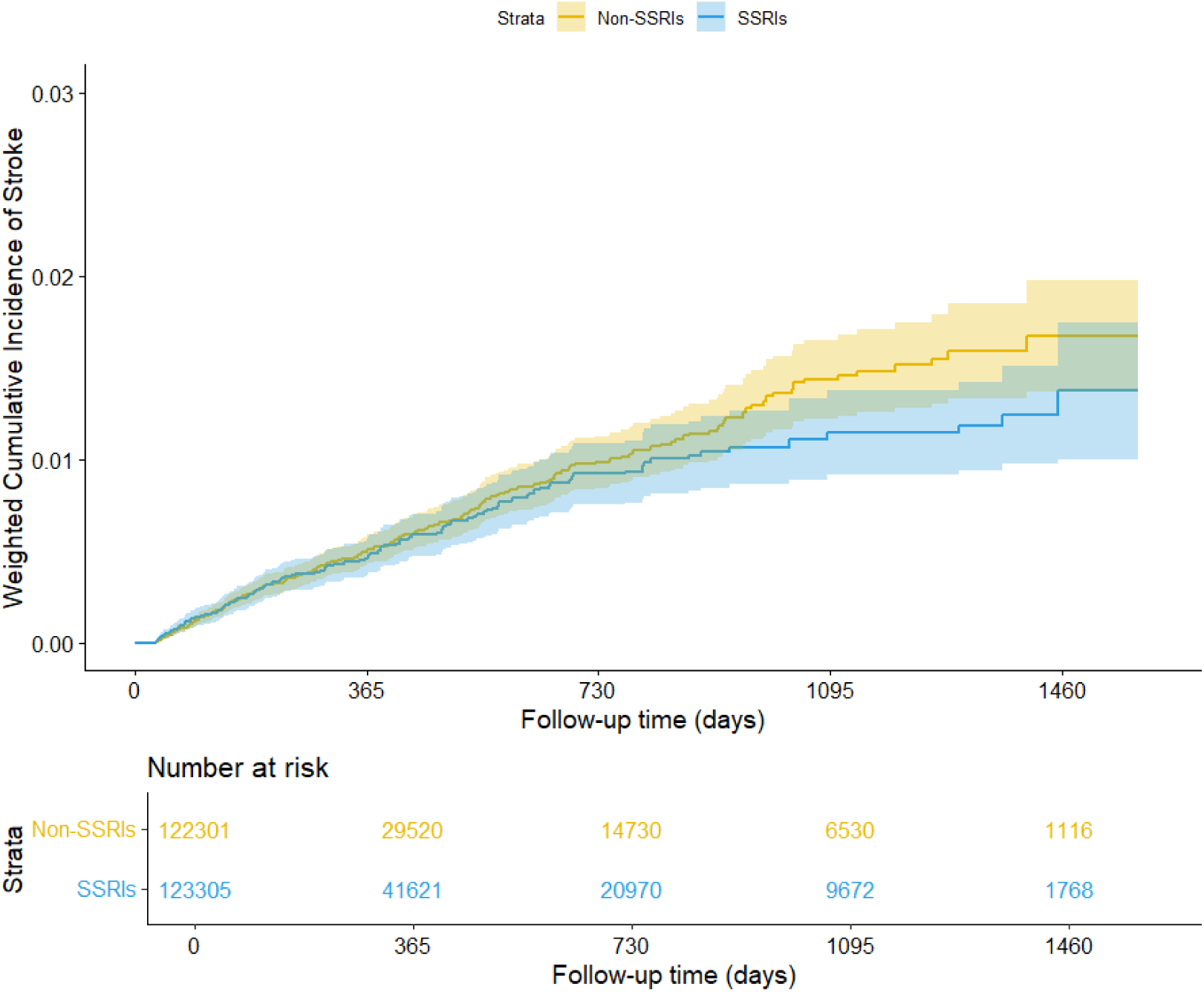
Weighted cumulative incidence of any type of stroke over follow-up period for SSRI or non-SSRI use. Yellow represents non-SSRI use and blue represents SSRI use. Number at risk represents the number of patients that remain within the cohort who had not been diagnosed of stroke or been censored at specific time intervals.

Results from doubly robust Cox model at each time point showed that SSRI users were associated with a non-significant lower risk of stroke compared to non-SSRI users (adjusted hazard ratio [AHR] 0.95; 95% CI 0.77 – 1.20), which is shown in **Table 2**. Further analysis consistently demonstrated that SSRI users were associated with a non-significant reduced risk of ischemic stroke (adjusted hazard ratio [AHR] 0.97; 95% CI 0.70 – 1.34) and hemorrhagic stroke (adjusted hazard ratio [AHR] 0.96; 95% CI 0.74 – 1.26) when compared to non-SSRI users in a doubly robust adjusted analysis (**Table 2**).

**Table 2.** Adjusted incidence rate ratio and hazard ratio of stroke.

| Outcomes |  | Cohort size | Average Follow-up days | Number of strokes | Crude incidence rate per 100,000 person-years | Weighted incidence rate per 100,000 person-years | Crude IRR with 95% CI | Weighted IRR with 95% CI | Doubly robust* *adjusted IRR* with 95% CI | Doubly robust* *adjusted HR* with 95% CI |
| --- | --- | --- | --- | --- | --- | --- | --- | --- | --- | --- |
| Any type of | Overall | 122,679 | 302 | 434 | 428.43 | 439.61 |  |  |  |  |
| stroke | SSRIs | 55,279 | 349 | 154 | 291.57 | 419.38 | 0.51 (0.42, 0.62) | 0.90 (0.73, 1.10) | 0.948 (0.77, 1.16) | 0.95 (0.77, 1.20) |
|  | Non-SSRIs | 67,400 | 263 | 280 | 577.52 | 466.61 | Ref. | Ref. | Ref. | Ref. |
| Ischemic stroke | Overall | 122,679 | 302 | 259 | 172.75 | 175.00 |  |  |  |  |
|  | SSRIs | 55,279 | 349 | 93 | 115.49 | 166.49 | 0.49 (0.36, 0.67) | 0.89 (0.65, 1.23) | 0.94 (0.68, 1.30) | 0.97 (0.70, 1.34) |
|  | Non-SSRIs | 67,400 | 263 | 166 | 235.13 | 186.36 | Ref. | Ref. | Ref. | Ref. |
| Hemorrhagic stroke | Overall | 122,679 | 302 | 175 | 255.67 | 264.61 |  |  |  |  |
|  | SSRIs | 55,279 | 349 | 61 | 176.08 | 252.90 | 0.51 (0.40, 0.66) | 0.90 (0.70, 1.17) | 0.95 (0.73, 1.24) | 0.96 (0.74, 1.26) |
|  | Non-SSRIs | 67,400 | 263 | 114 | 342.39 | 280.26 | Ref. | Ref. | Ref. | Ref. |

\*: Doubly robust means after using inverse probability weight (IPW) to balance the baseline characteristics distribution between SSRIs and non-SSRIs group, we adjusted these confounders in the final model with IPW as well, since the SMD shows an imbalanced distribution of confounders after IPW.

**Fig. 3** showed that the incidence of ischemic stroke among SSRI users is slightly lower than that among non-SSRI users throughout the entire follow-up time, while **Fig. 4** showed that the incidence of hemorrhagic stroke among SSRI users is nearly indistinguishable from that among non-SSRI users.

**Fig. 3.**
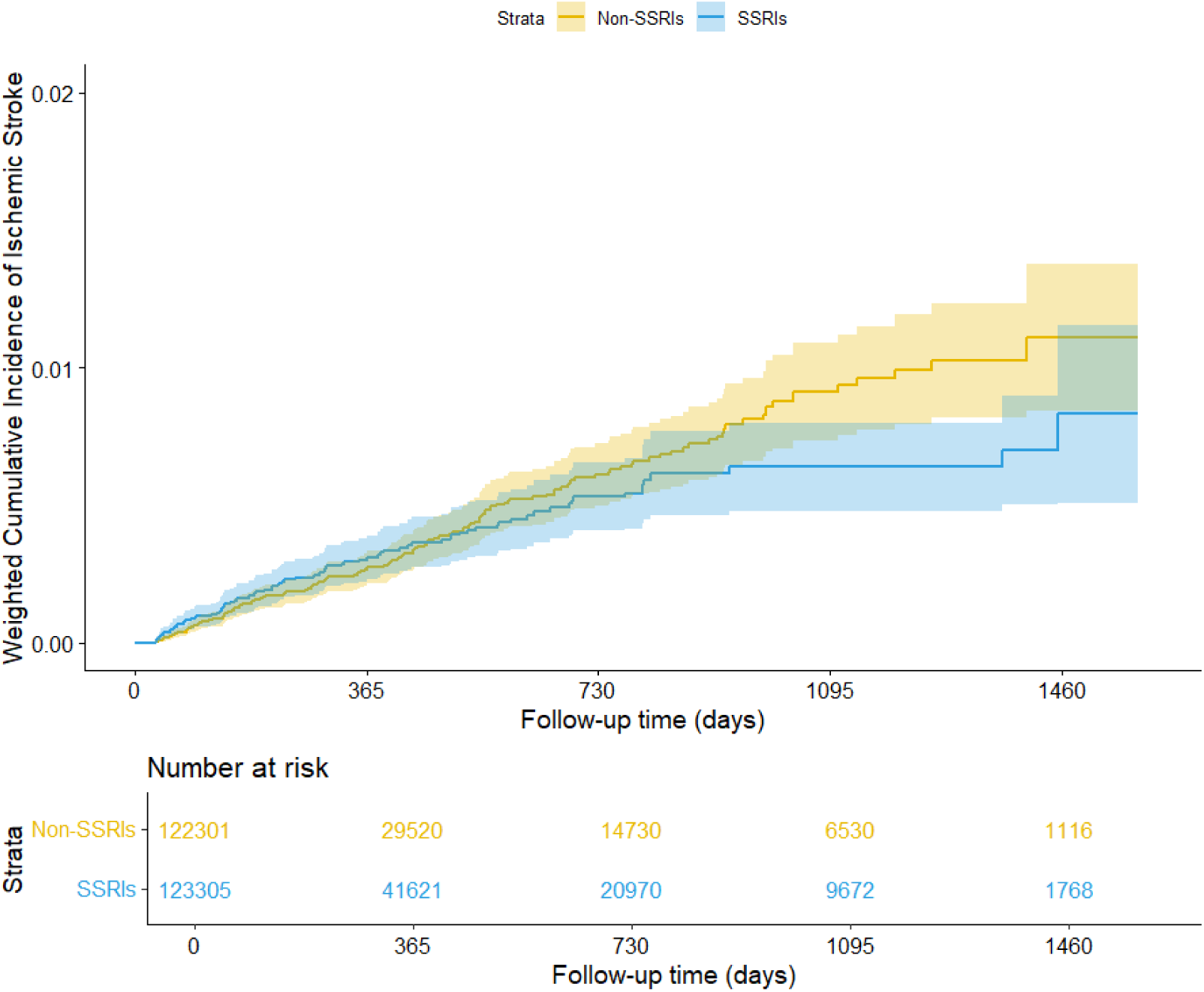
Weighted cumulative incidence of ischemic stroke over follow-up period for SSRI or non-SSRI use. Yellow represents non-SSRI use and blue represents SSRI use. Number at risk represents the number of patients that remain within the cohort who had not been diagnosed of ischemic stroke or been censored at specific time intervals.

**Fig. 4.**
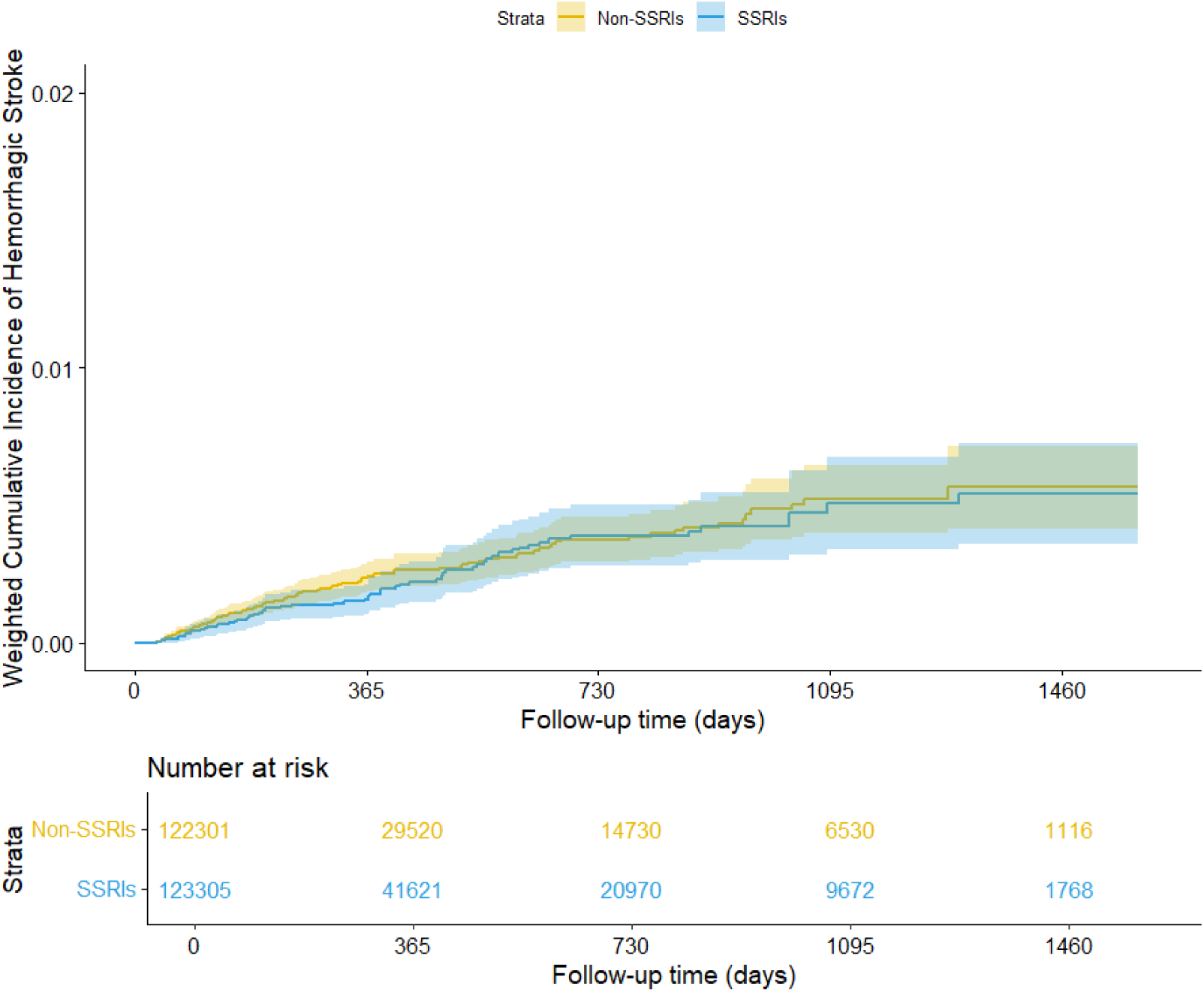
Weighted cumulative incidence of hemorrhagic stroke over follow-up period for SSRI or non-SSRI use. Yellow represents non-SSRI use and blue represents SSRI use. Number at risk represents the number of patients that remain within the cohort who had not been diagnosed of hemorrhagic stroke or been censored at specific time intervals.

### Systematic Review

As shown in **Fig. 5**, 813 articles were retrieved from online database upon initial search, of which 191 duplicates were removed. 575 articles were further excluded during title and abstract screening process. Upon a comprehensive evaluation of eligibility of the remaining 47 articles through full-text screening, 5 observational studies were included in quality assessment [6, 10, 21–23]. The inter-rater reliability of observational studies screening process was 0.96, underscoring a robust agreement between the two investigators involved. 5 observational studies, together with newly conducted cohort study provided adequate data for pooled estimates of hypothesized associations. Study features, quality appraisal and study results are listed in **Tables 3 and 4**.

**Fig. 5.**
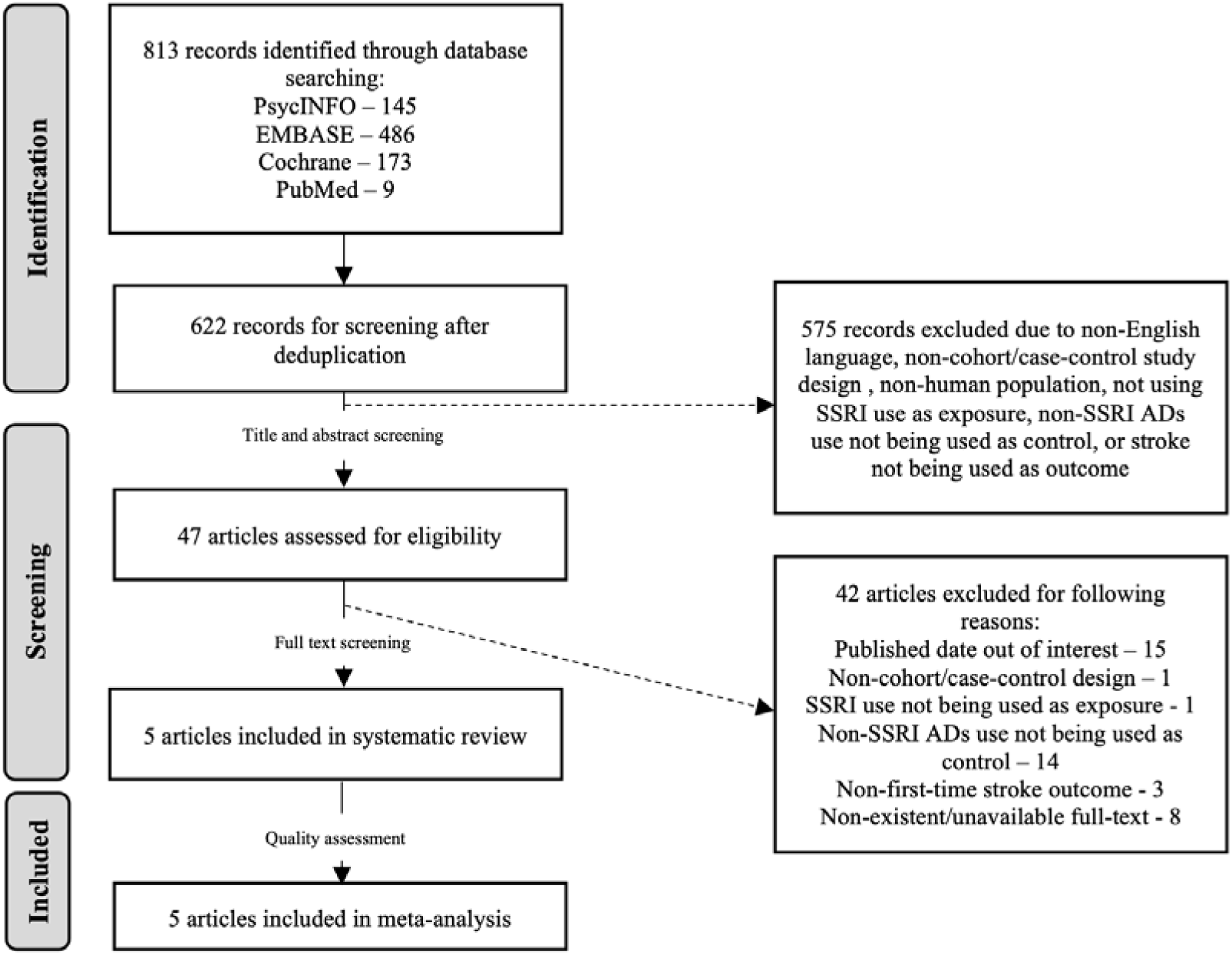
Flow chart of study selection

**Table 3.** Characteristics and results of quality appraisal of included studies.

| Study | Region | Data source | Study design | Sample size | Inclusion criteria | Exclusion criteria | Outcome definition | NOS Score |  |  |
| --- | --- | --- | --- | --- | --- | --- | --- | --- | --- | --- |
|  |  |  |  |  |  |  |  | Selection | Comparability | Exposure/<br>Outcome |
| Douros *et al.* (2019) [6] | United Kingdom | UK Clinical Practice Research Datalink (CPRD) | Nested case control study | 938,388<br>Cases: 15,860<br>Controls: 473,712 | age ≥18 years and newly prescribed an SSRI or a 3<sup>rd</sup> generation AD between January 1, 1995, and June 30, 2014 | <1 year of information in the CPRD, being prescribed AD before cohort entry, use of trazodone / MAOI / TCA, MI / stroke history | Ischemic stroke / TIA | **** | ** | ** |
| Alqdwhah-Fattouh *et al.* (2022) [10] | Spain | BIFAP | Nested case control study | 3,757,621<br>Cases: 8296<br>Controls: 37,252 | 40 – 99 years old without any previous record of cancer or stroke, at least 1-year registry with physician over the study period | Stroke with cardioembolic origin, strokes of unusual origin, recorded prescriptions of any anti-depressant before start date) | Non-cardioembolic ischemic stroke | **** | ** | ** |
| Albrecht *et al.* (2018) [21] | United States | CMS chronic condition data warehouse | Cohort study | 30,886 | age ≥ 65 years Medicare beneficiaries hospitalized with first TBI during 2006–2010, and survival to hospital discharge | First 2 months post-TBI | Ischemic stroke/<br>Hemorrhagic stroke | ** | ** | ** |
| Almuwaqqat *et al.* (2019) [22] | United States | ARIS study center | Cohort study | SSRI exposed: 944<br>Non-SSRI AD exposed: 1083 | 45 – 64 years old participated in any of the first 5 visits and reported use of ADs at any of the visits | MI / HF / stroke / AF history, recorded ADs medication use history, non-white subjects from Minnesota and Maryland centers | Ischemic stroke | ** | ** | ** |
| Renoux *et al.* (2017) [23] | United Kingdom | UK Clinical Practice Research Datalink (CPRD) | Nested case control study | 1,363,990<br>Cases: 3036<br>Controls: 89,702 | age ≥18 years and newly prescribed an AD between January 1, 1995 and June 30, 2014 | <1 year of information in the CPRD before cohort entry, been dispensed any ADs, history of stroke (ischemic or hemorrhagic) or TIA | ICH | **** | ** | ** |

BIFAP, Pharmacoepidemiological Research Database for Public Health Systems; CMS, Centers for Medicare & Medicaid Services; ARIS, Atherosclerosis Risk in Communities

**Table 4.** Results of 5 included studies.

| Study | Sample size | Exposed group definition | Number of cases in 'exposed group' | Association between SSRI use and stroke using non-SSRI as comparator (95% CI) | Adjusted covariates |
| --- | --- | --- | --- | --- | --- |
| Douros *et al.* (2019) [6] | Cases: 15,860<br>Controls: 473,712 | Prescription for SSRI or a third-generation antidepressant | Strong inhibitors of serotonin reuptake: 2,277<br>Weak inhibitors of serotonin reuptake: 559 | SSRI ARR: 0.88 (0.80-0.97)<br>(weak inhibitors of serotonin reuptake as comparator) | Age, sex, duration of follow up, calendar time, obesity, smoking status, alcohol abuse, arterial hypertension, atrial fibrillation, congestive heart failure, myocardial infarction, other coronary artery disease, peripheral vascular disease, hyperlipidaemia, diabetes mellitus, chronic kidney disease, liver disease, depression chronic obstructive pulmonary disease, cancer, use of anticoagulants, antiplatelets, lipid-lowering drugs, nonsteroidal anti-inflammatory drugs, opioids B-blockers, thiazides, angiotensin-converting enzyme inhibitors, angiotensin II receptor blockers, calcium channel blockers, antipsychotics, other anti-depressants (tricyclic antidepressants, monoamine oxidase inhibitors, trazodone), lithium, hormone replacement therapy, and number of physician visits |
| Alqdwhah-Fattouh *et al.* (2022) [10] | Cases: 8296<br>Controls: 37,252 | Recorded prescriptions for antidepressants | SSRI: 255<br>Non-SSRI-nonSARI: 175 | SSRI AOR: 0.74 (0.58-0.93)<br>*Converted RR: 0.80 (0.66-0.95)* | Age, sex, year, Visits (last 12 month), BMI, Smoking, Alcohol abuse, Ischemic heart disease, Heart failure, TIA, PAD, Hypertension, Diabetes, Dyslipidaemia, COPD, Rheumatoid arthritis, Chronic kidney failure, Hyperuricemia, (Current use of Antiplatelet drugs, Paracetamol, Metamizole, NSAIDs, Opioids, Corticosteroids, ACEIs, ARBs, CCBs, Beta-blockers, Alfa-blockers, Diuretics, PPIs, H2-receptor antagonists, Benzodiazepines, Antipsychotics) |
| Albrecht *et al.* (2018) [21] | Participants: 30,886 | Prescription for SSRI, SNRI, and TCA | *Haemorrhagic stroke*<br>SSRI: 431<br>SNRI: 96<br>TCA: 41<br>*Ischemic stroke*<br>SSRI: 2142<br>SNRI: 466<br>TCA: 243 | *Haemorrhagic stroke*<br>SSRI ARR: 1.17 (0.71-0.92)<br>(SNRI as comparator)<br>SSRI ARR: 2.47 (1.30-4.70)<br>(TCA as comparator)<br>*Ischemic stroke*<br>SSRI ARR: 0.92 (0.73-1.15)<br>(SNRI as comparator)<br>SSRI ARR: 0.99 (0.76-1.31)<br>(TCA as comparator) | *Haemorrhagic stroke*<br>Age, sex, race, atrial fibrillation, heart failure, chronic obstructive pulmonary disease (COPD), hip fracture, osteoarthritis, stroke, pre-TBI use of each of the compared classes<br>*Ischemic stroke*<br>Age, sex, race, atrial fibrillation, heart failure, diabetes, liver disease, neurologic disease, hip fracture, hypertension, ischemic heart disease, stroke, pre-TBI use of each of the compared classes |

**Table 4.** Results of 5 included studies (continued)
| Study | Sample size | Exposed group definition | Number of cases in 'exposed group' | Association between SSRI use and stroke using non-SSRI as comparator (95% CI) | Adjusted covariates |
| --- | --- | --- | --- | --- | --- |
| Almuwaqq at *et al.* (2019) [22] | Exposed to SSRI: 944<br>Exposed to non-SSRI: 1083 | Antidepressant medications brought to visit | SSRI: 43<br>Non-SSRI: 76 | SSRI AHR: 0.91 (0.62-1.35)<br>*Converted RR: 0.87 (0.47–1.57)* | BMI, cigarette smoking, alcohol use, antihypertensive medications, diabetes mellitus (HDL, LDL, statin use for MI and stroke models), year of diagnosis |
| Renoux *et al.* (2017) [23] | Cases: 3036<br>Controls: 89,702 | Prescription for antidepressant from computerized medical records | SSRI: 588<br>TCA: 364 | SSRI ARR: 1.17 (1.02-1.35) | Age, Male sex, Body mass index, Smoking status, Alcohol abuse, Hyperlipidaemia, Hypertension, Diabetes, Transient ischemic attack, Atrial fibrillation, Coronary artery disease, Congestive heart failure, Peripheral vascular disease, Chronic obstructive pulmonary disease, Renal failure, Depression, Cancer, Liver disease, Disorders of haemostasis, Brain vascular disease, History of bleeding, No. of physician visits, Anticoagulant use, Antiplatelet use, NSAID use, Antipsychotic use, pain |

Abbreviations: AHR, adjusted hazard ratio; AOR, adjusted odd ratio; ARR, adjusted risk ratio; ARBs, Angiotensin receptor blockers; ACEIs, Angiotensin-converting enzyme inhibitors; CCBs, Calcium channel blockers; H2, Histamine-2; LDL, Low-density lipoprotein; MI, Myocardial Infarction; NSAID, Nonsteroidal Anti-Inflammatory Drug; PPIs, Proton pump inhibitors; SARI, Serotonin antagonist and reuptake inhibitor; SSRI, selective serotonin reuptake inhibitor; TBI, Traumatic Brain Injury; TCA, Tricyclic antidepressants

#### Study characteristics

The studies incorporated in the analysis were carried out in United States [21, 22], United Kingdom [6, 23], and Spain [10]. Among these, three were nested case-control studies [6, 10, 23], while two were cohort studies [21, 22]. The sample sizes varied across the studies, ranging from 2027 [22] to over 400,000 individuals [6]. All studies received a moderate to high rating in quality assessment, with NOS score between 6-8 stars.

#### Exposure – SSRI and non-SSRI use

All studies used electronic database records to identify the use of SSRI and non-SSRI except for Almuwaqqat et al., which delineated the exposed group based on antidepressants physically brought to clinical visits by study participants [22]. Exposure durations were specified in two studies, Douros et al. incorporated individuals exposed to the antidepressant of interest (SSRI or a third-generation antidepressant) within 30 days before the diagnosis of ischemic stroke [6]; Renoux et al. included individuals exposed to antidepressants if the duration of their most recent prescription included the date of the intracranial hemorrhage diagnosis or ended within 30 days prior [23].

Out of the 5 studies, 4 studies assessed the risk of ischemic stroke associated with SSRI exposure [6, 10, 21, 22]. Of those, 2 reported no association between SSRI use and ischemic stroke [21, 22], with Albrecht et al. reported an adjusted risk ratio (ARR) of 0.92 (95% CI 0.73-1.15) and ARR 0.99 (95% CI 0.76-1.31) using SNRI and TCA as comparator respectively, while Almuwaqqat et al. indicated adjusted hazard ratio (AHR) of 0.91 (95% CI 0.62-1.35). A reduction in ischemic stroke risk was suggested 2 another studies [6, 10]. Specifically, Douros et al. classified antidepressants according to their dissociation constant (K_D_), with the category of strong inhibitors of serotonin reuptake only composed of SSRIs and no SSRI was grouped into the category of weak inhibitors, and they showed a decreased risk of ischemic stroke with the use of strong inhibitors of serotonin reuptake (ARR 0.88; 95% CI 0.80 – 0.97) [6]. Alqdwah-Fattouh et al. showed similar decreases in ischemic stroke risk (adjusted odd ratio [AOR] 0.74; 95% CI 0.58 – 0.93) [10].

Two studies investigated the risk of hemorrhagic stroke associated with SSRI exposure [21, 23]. Of those, Albrecht et al. found that exposure to SSRI were strongly associated with an increased risk of hemorrhagic stroke when compared with TCA (ARR 2.47; 95% CI 1.30 – 4.70) [21]. Yet, this association was reported statistically insignificant when SNRI was used as comparator (ARR 1.17, 95% CI 0.71 – 1.92) [21]. Renoux et al. also found a small association between hemorrhagic stroke risk and exposure to SSRI when compared with TCA (ARR 1.17; 95% CI 1.02 – 1.35) [13].

Some studies have also investigated the duration of SSRI exposure and its association with stroke [6, 23], with Douros et al. revealed that the potential protective effects of SSRI exposure appeared to be more pronounced within a specific timeframe lasting between 61 to 180 days, demonstrating ARR of 0.84 (95% CI 0.72 – 0.97). Renoux et al. stratified the duration of SSRI exposure into three distinct intervals: ≤30 days, 31-90 days, and >90 days, and they reported the risk of hemorrhagic stroke was higher when SSRI was used for not more than 30 days compared with TCA (ARR 1.44; 95% CI 1.04 – 1.99) [23].

#### Outcome

All five studies defined the outcome of interest as the first diagnosis of stroke, with three studies explicitly indicating the diagnostic criteria based on International Classification of Diseases code (ICD) [10, 22, 22]. In addition to using ICD codes for diagnosis, two other studies defined diagnosis using Read code [6, 23], which is a clinical terminology system widely used in UK [24].

#### Confounder adjustment

The potential confounder addressed in the five studies can be classified into four primary domains: sociodemographic factors, lifestyle variables, baseline comorbidities, and medical history.

The major sociodemographic factors adjusted include age, sex, race and education level. Four studies also adjusted for lifestyle factors, including BMI, smoking, and alcohol use [6, 10, 22, 23]. All studies identified comorbidities and medical history as potential confounders and adjusted accordingly. 4 studies [6, 10, 22, 23] have adjusted the use of medications which have been identified to affect stroke risk, such as antithrombotic agents and lipid lowering agents [25, 26]. One study adjusted for vital exhaustion questionnaire, which vital exhaustion has been established as a risk factor for cardiovascular events [27].

#### Quality assessment

All studies have received six to eight stars in the NOS quality assessment according to the procedures mentioned in **appendix 4**. One cohort study received a low rating due to inadequate description regarding the selection procedure and sample size of exposed individual [21]. Another cohort study has a reduced rating due to a limited sample size of exposed individuals and uncertain ascertainment of exposure [22].

#### Pooled estimates of the association

No articles were excluded from the meta-analysis. To ensure the effect measures used to report associations were comparable among the studies, an online calculator was used to convert odd ratio to risk ratio [28]. Hazard ratio was converted to risk ratio using online calculator with the following formula (HR=fracln(OR)1.65) [29]. Using a random effect model, we pooled the RRs of stroke risk between SSRI users and non-SSRI users from all 5 observational studies, together with the retrospective cohort study conducted, with the I^2^ estimated at 63%. **Fig. 6** shows the forest plot result and pooled measure of association.

**Fig. 6.**
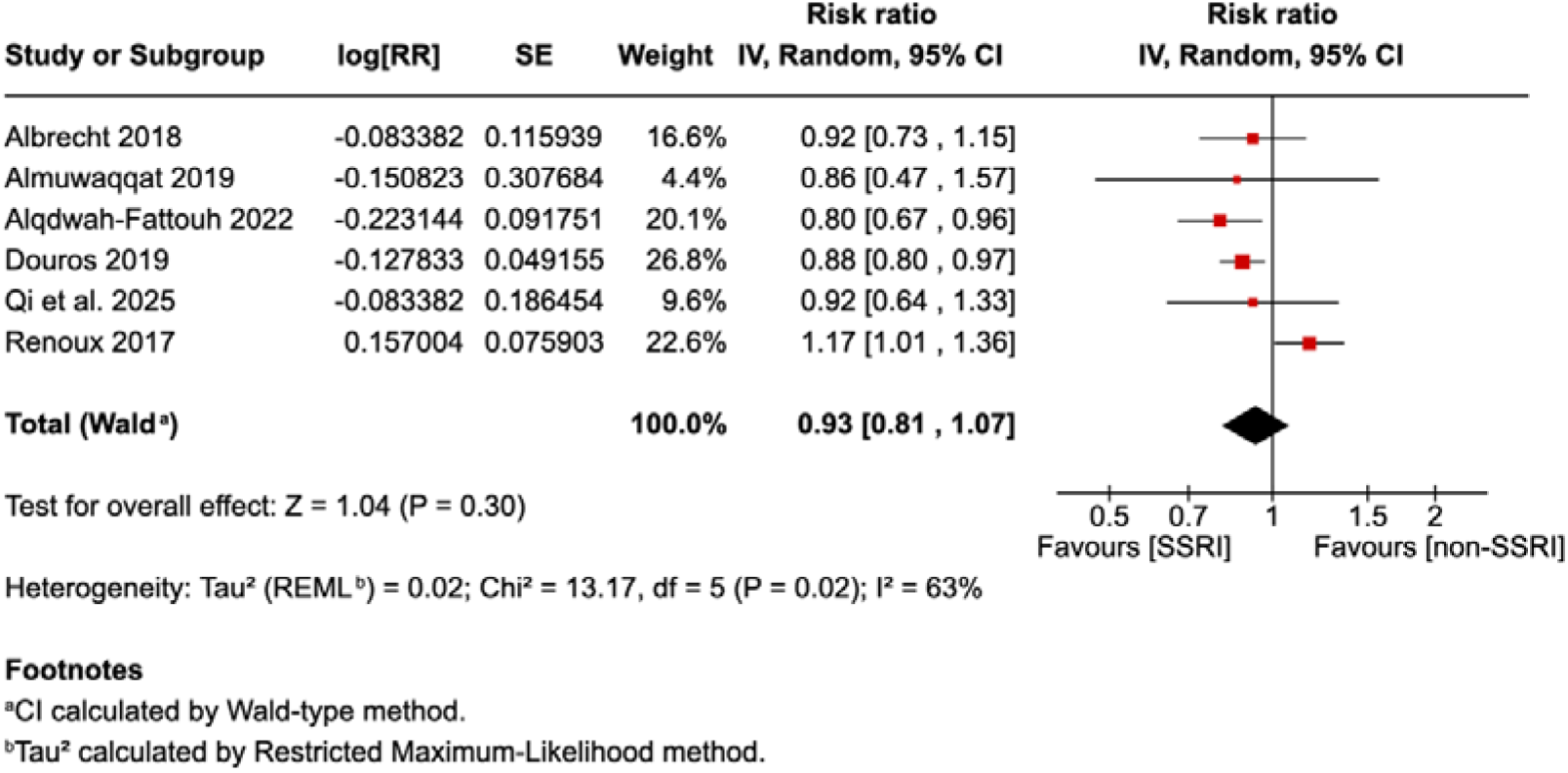
Forest plot showing RRs generated from retrieved observational studies (*n* = 5) and newly conducted retrospective cohort study.

Result indicates SSRIs (as compared with non-SSRIs) were not associated with statistically different risk of stroke (RR 0.93, 95% CI 0.81-1.07). Due to limited inclusion of studies in the pooled estimates, we did not perform Egger’s regression test to assess publication bias.

As one of the cohort studies [21] stratified the analysis by using TCA/SNRI as comparator and hemorrhagic/ischemic stroke as outcome, we incorporated the RR using SNRI as comparator and ischemic stroke as outcome. The analysis was subsequently replicated with all three others RR separately as a sensitivity measure to assess result robustness. No substantial difference was observed, as displayed in **appendix 5**.

## DISCUSSION

Compared with previous studies that did not use an active-comparator design, findings from our large cohort study involving approximately 100,000 individuals initiating antidepressant treatment suggested that SSRI use is potentially associated with a lower risk of stroke compared to non-SSRI antidepressants. Proper adjustments for potential confounders, such as the use of antiplatelet and antithrombotic agents and underlying cardiovascular diseases, were made. Subgroup analyses focusing on stroke subtypes (i.e. ischemic and hemorrhagic strokes) consistently demonstrated a similar statistically non-significant association. Results of the systematic review and meta-analysis did not support any association between SSRI use and an increased risk of stroke compared to non-SSRI antidepressants.

To the best of our knowledge, there were only two cohort studies investigating the association between SSRI use and stroke risk using non-SSRI antidepressants as comparator [21, 22]. Of these, one of the studies used individuals hospitalized with traumatic brain injury (TBI) as study population [21]. This may introduce selection bias into the study given, as recent meta-analysis indicated TBI is associated with a substantial increase in stroke risk [30], thereby potentially limiting the external validity of the study findings [21]. Additionally, the other study had an extensive follow-up period exceeding 25 years, yet the analysis solely adjusted for baseline covariates [22]. Given the time-varying nature of covariates, such as age and disease status, this approach may introduce error to the study findings. Consequently, the retrospective cohort study was conducted to address these gaps in existing literature, with an average follow-up duration of approximately a year.

With no randomization performed, there remains to be a risk of confounding by indication: the observed association between SSRI use and stroke may only stem from an underlying factor that cause both SSRI prescription and the onset of stroke. Thus, our study design highlights the importance for future studies to employ an active comparator approach. This may also explain the difference in findings between our meta-analysis and previous studies which indicated that SSRI use was significantly associated with an increase in stroke risk [2, 31]. This method is crucial for minimizing the influence of potential indication bias, thereby strengthening the reliability and validity of findings in uncovering the true association between SSRI use and the risk of stroke. It is noteworthy that all studies included in the meta-analysis adhered to this method.

### Strengths and limitations

Our cohort study used electronic health records from a large territory-wide database for analysis. This allowed us to have a large sample size with real clinical data, making our findings more accurate and representative of the population. Moreover, we included only observational studies that utilized active comparators in our review. This approach helped reduce the influence of confounding by indication in our analysis.

In spite of its potential clinical implications, the study has several limitations. First, exposure was ascertained by prescription records in four of the studies, including our cohort study [6, 10, 21, 23]. Yet, prescription records do not provide insights into whether individuals effectively followed their SSRI medication regimens due to common non-adherence to SSRIs, often driven by side effects such as sexual dysfunction and somnolence [32, 33]. A recent study indicated that about 20% of users did not adhere to SSRI treatment [32]. This non-adherence poses a risk of bias in study findings. Secondly, all studies included in the meta-analysis relied on electronic databases as their primary data source. This approach carries inherent limitations, including the possibility of neglecting manually documented clinical histories when they are not incorporated into the electronic database, as well as a lack of comprehensive information regarding various confounding factors like lifestyle variables, thereby posing challenges in adjusting these covariates during subsequent analysis. Consequently, these limitations have the potential to introduce bias into the study findings. Furthermore, only observational studies without randomization were reviewed and included in meta-analysis, in which unmeasured confounding effects were not able to match between the exposed and nonexposed cohort. This may introduce confounding bias to our study findings.

## CONCLUSION

In summary, our analysis revealed no association between SSRI use and stroke risk, with our original cohort study showing a potentially lower risk compared with non-SSRI antidepressants. However, given the considerable inter-study variability in results, further investigation is necessary to validate the null association.

## Data Availability

Data will not be accessible to others as the data custodian has not granted permission.

## Abbreviations and Acronyms

SSRI: selective serotonin reuptake inhibitor
SNRI: serotonin and norepinephrine reuptake inhibitor
AD: antidepressant
TCA: tricyclic antidepressant
MAOI: monoamine oxidase inhibitor
MI: myocardial infarction
TBI: traumatic brain injury
HF: heart failure
AF: atrial fibrillation
TIA: transient ischemic attack
ICH: intracranial hemorrhage

## Contributors’ Statement

Paco Wong Chun Yeung and Francisco Lai contributed the conception of this work. The study was designed by all authors. Paco Wong Chun Yeung and Qi Sun contributed to the acquisition and analysis of the data for systematic review and cohort study, respectively, and all authors interpreted the data. Paco Wong Chun Yeung, Qi Sun, and Francisco Lai drafted the manuscript. Every contributor critically reviewed the manuscript for essential intellectual content, endorsed the final version for publication, and accepted responsibility for all components of the work. Paco Wong Chun Yeung and Qi Sun are the co-first authors and Francisco Lai is the corresponding author. Francisco Lai had full access to all the data in this work and all authors took ultimate responsibility for the decision to submit this manuscript for publication.

## Acknowledgements

The authors thank the Hospital Authority for the generous provision of data.

## Appendix 1

| *Comorbidities history* | ICD-9 codes |
| --- | --- |
| hypertension | 401 |
| atrial fibrillation | 427.31 |
| diabetes | 250 |
| hyperlipidemia | 272.0-272.4 |
| obesity | 278.00, 278.01 |
| chronic kidney disease | 585.1-585.6, 585.9 |
| obstructive sleep apnea | 327.23 |
| coronary artery disease | 414.00 |
| heart failure | 428.0 |
| carotid artery stenosis | 433.10 |
| tobacco use disorder | 305.1 |
| alcohol abuse | 305.00 |
| migraine with aura | 346.00, 346.01 |
| sickle cell disease | 282.60 |
| systemic lupus erythematosus | 710.0 |
| rheumatoid arthritis | 714.0 |
| antiphospholipid syndrome | 289.81 |
| major depressive disorder | 296.20-296.26, 296.30-296.36 |
| anxiety disorders | 300.00 |
| peripheral artery disease | 443.9 |
| human immunodeficiency virus | 042 |
| lung cancer | 162 |
| pancreatic cancer | 157 |
| gastrointestinal cancers | 151, 153, 154 |
| hematologic malignancies | 200.0-200.3; 202; 202.8; 204-208 |
| brain tumors | 191, 198.3 |
| head and neck cancers | 140.0, 141.0, 143.0, 146.0, 148.0 |
| cocaine abuse | 305.60 |
| amphetamine abuse | 305.70 |
| chronic obstructive pulmonary disease | 496 |
| sepsis | 038 |
| *Stroke type* | ICD-9 codes |
| Ischemic Stroke | 434 |
| Hemorrhagic Stroke | 430, 431 |
| *Medication usage within one year before the index date* | B.N.F. code |
| warfarin | 2.8.2 |
| dabigatran | 2.8.2 |
| rivaroxaban | 2.8.2 |
| nonsteroidal anti-inflammatory drugs | 10.1.1 |
| oral contraceptives | 7.3.1 |
| estrogens | 6.4.1.1 |
| antiplatelet agents | 2.9 |
| decongestants | 3.10 |
| atypical antipsychotics | 4.2.1 |
| erythropoiesis-stimulating agents | 9.1.3 |
| antihypertensives | 2.5.5.1, 2.5.5.2, 2.4, 2.6.2 |
| statins | 2.12 |
| antiplatelet agents | 2.9 |
| antidiabetic medications | 6.1.2.2, 6.1.2.3, 6.1.2.5 |
| varenicline | 4.10.2 |

## Appendix 2 (keywords)

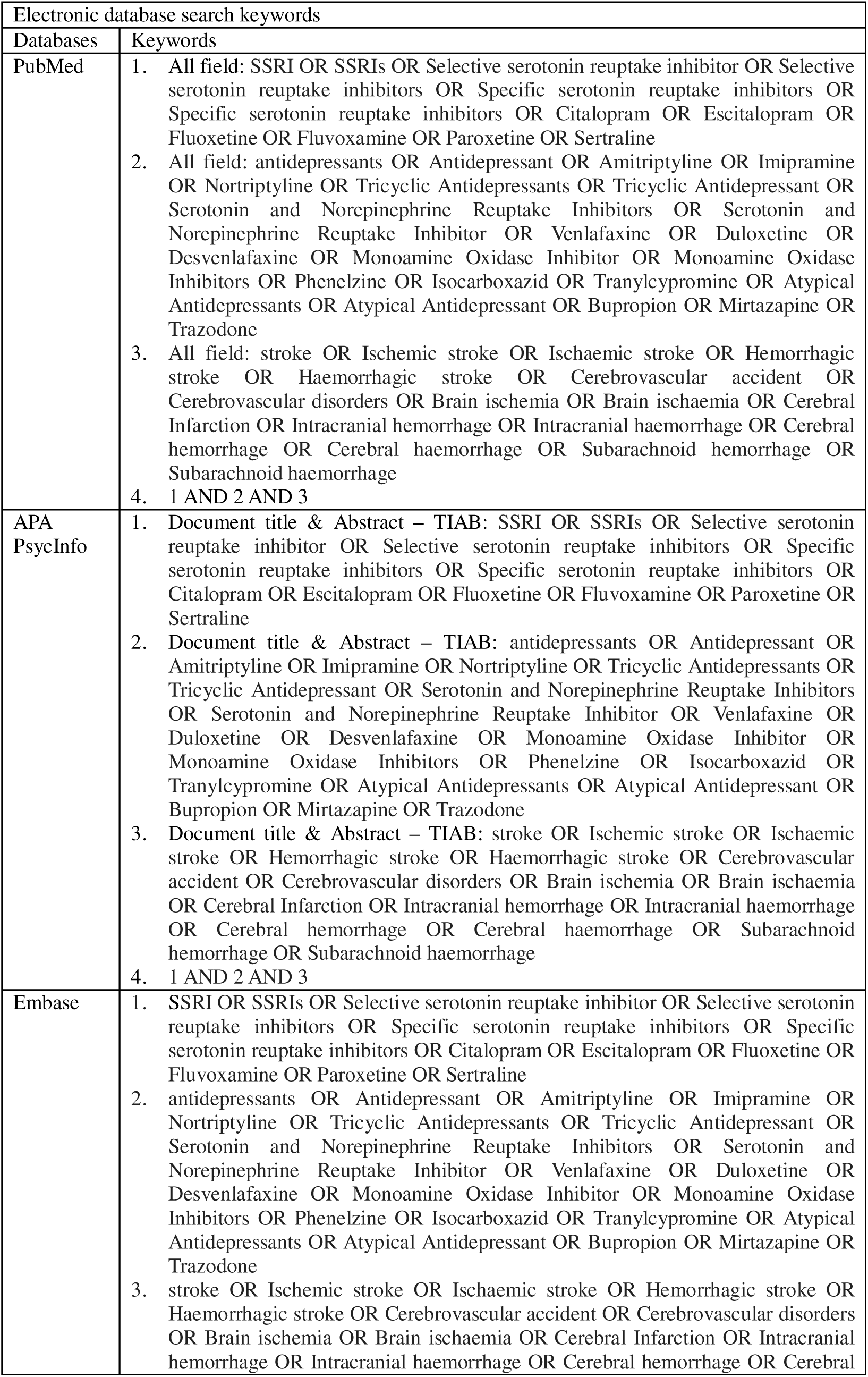
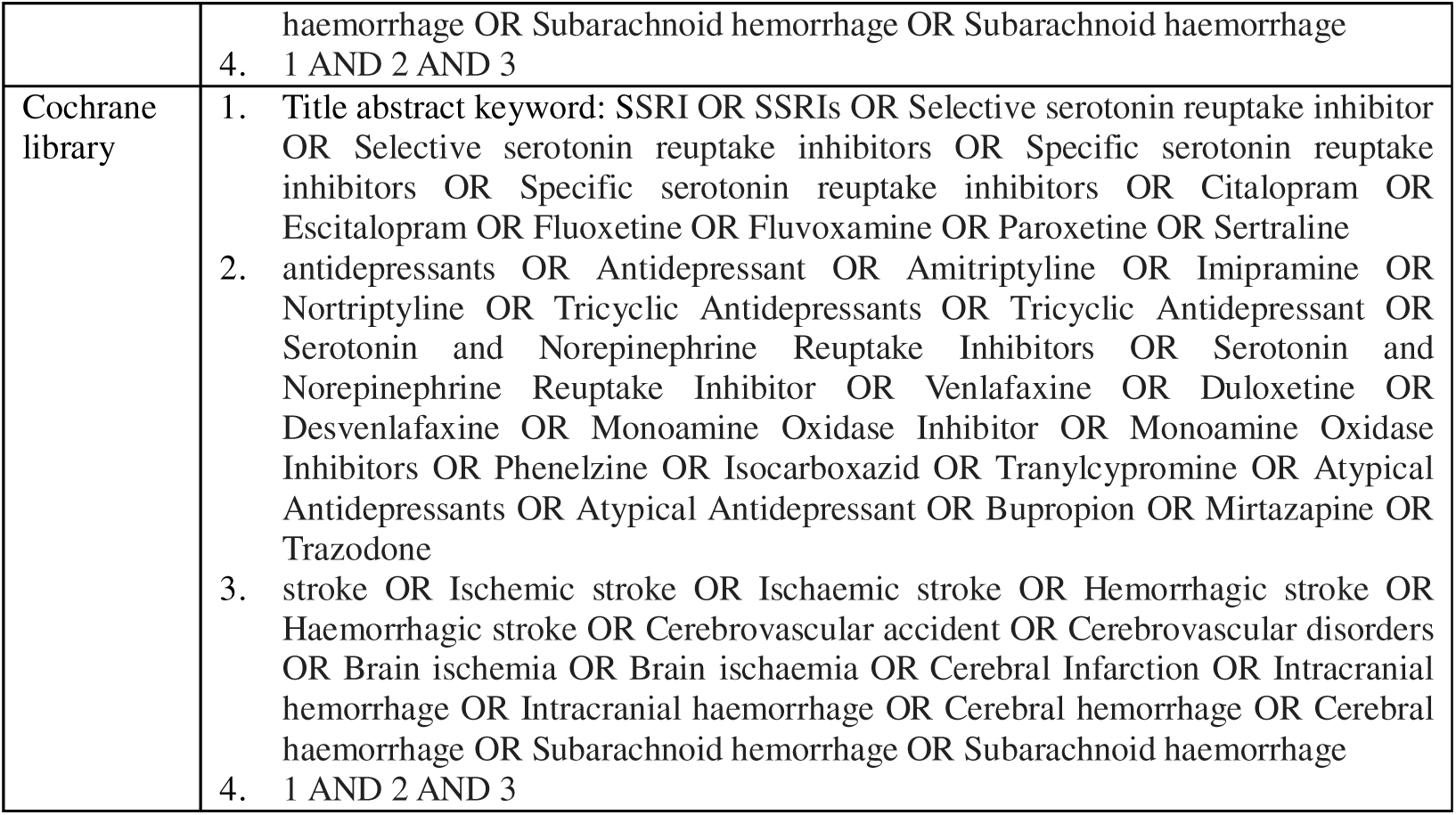

**Date of search: 9/8/2024**

**PsycINFO: 145 articles found**

**EMBASE: 486 articles found**

**Cochrane: 173 articles found**

**PubMed: 9 articles found**

## Appendix 3: Eligibility

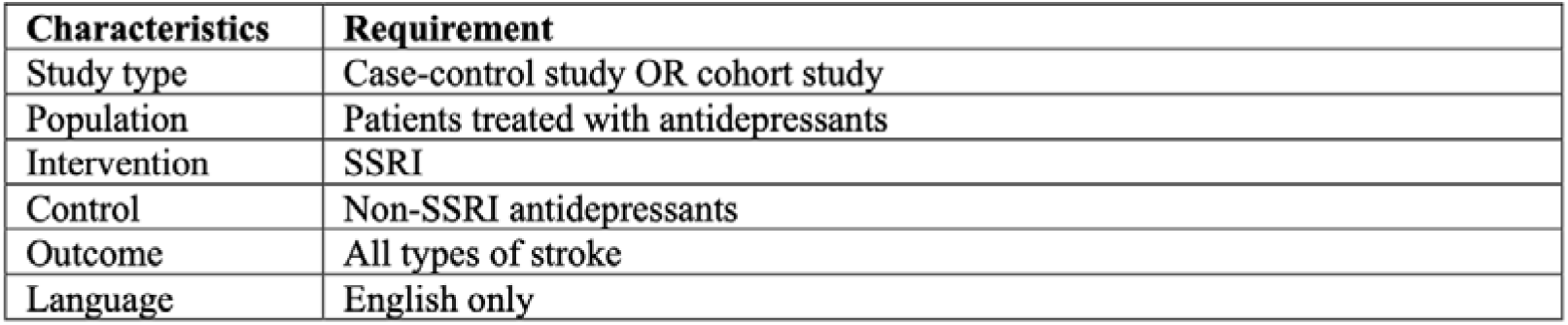

## Appendix 4: Newcastle Ottawa quality assessment scale

| Section | Case-control studies | Cohort studies |
| --- | --- | --- |
| Selection | 1. Case definition:<br>Allocate one star if cases are defined with independent validation (e.g. diagnostic code) | 1. Representativeness of exposed cohort:<br>Allocate one star if the sample size of exposed individuals > 1000 |
|  | 2. Representativeness of cases:<br>Allocate one star if sample size > 1000 | 2. Selection of the non-exposed cohort:<br>Allocate one star if non-exposed cohort were selected from the same community as the exposed cohort |
|  | 3. Selection of controls:<br>Allocate one star if community controls were used | 3. Ascertainment of exposure:<br>Allocate one star if exposure was ascertained by secure records (e.g. prescription records) OR by interview |
|  | 4. Definition of controls:<br>Allocate one star if controls had no stroke history (ischemic stroke / hemorrhagic stroke) | 4. Demonstration that outcome of interest was not present at start of study:<br>Allocate one star if all participants did not have stroke history before study entry. |
| Comparability | 5. Study controls for most important factor:<br>Allocate one star if study controls for clinical history | 5. Study controls for most important factor:<br>Allocate one star if study controls for clinical history |
|  | 6. Study controls for additional factors:<br>Allocate one star if study controls for factors other than clinical history (e.g. concurrent medications, sociodemographic) | 6. Study controls for additional factors:<br>Allocate one star if study controls for factors other than clinical history (e.g. concurrent medications, sociodemographic) |
| Exposure<br>(Case-control)/<br>Outcome<br>(Cohort) | 7. Ascertainment of exposure:<br>Allocate one star if exposure was ascertained by secure records (e.g. prescription records) OR by interview which is blind to case/control status | 7. Assessment of outcome:<br>Allocate one star if outcome is defined with independent assessment OR diagnostic record (e.g. ICD) |
|  | 8. Same method of ascertainment for cases and controls:<br>Allocate one star if the same method of ascertainment were used | 8. Was follow-up long enough for outcomes to occur:<br>Allocate one star if follow-up period > 3 years |
|  | 9. Non-Response rate:<br>Allocate one star if non-response rate was mentioned | 9. Adequacy of follow up of cohorts:<br>Allocate one star if complete follow up OR < 20% loss during follow up |

## Appendix 5: Forest plot of 5 observational studies RRs for SSRI use and stroke risk

**Fig.**
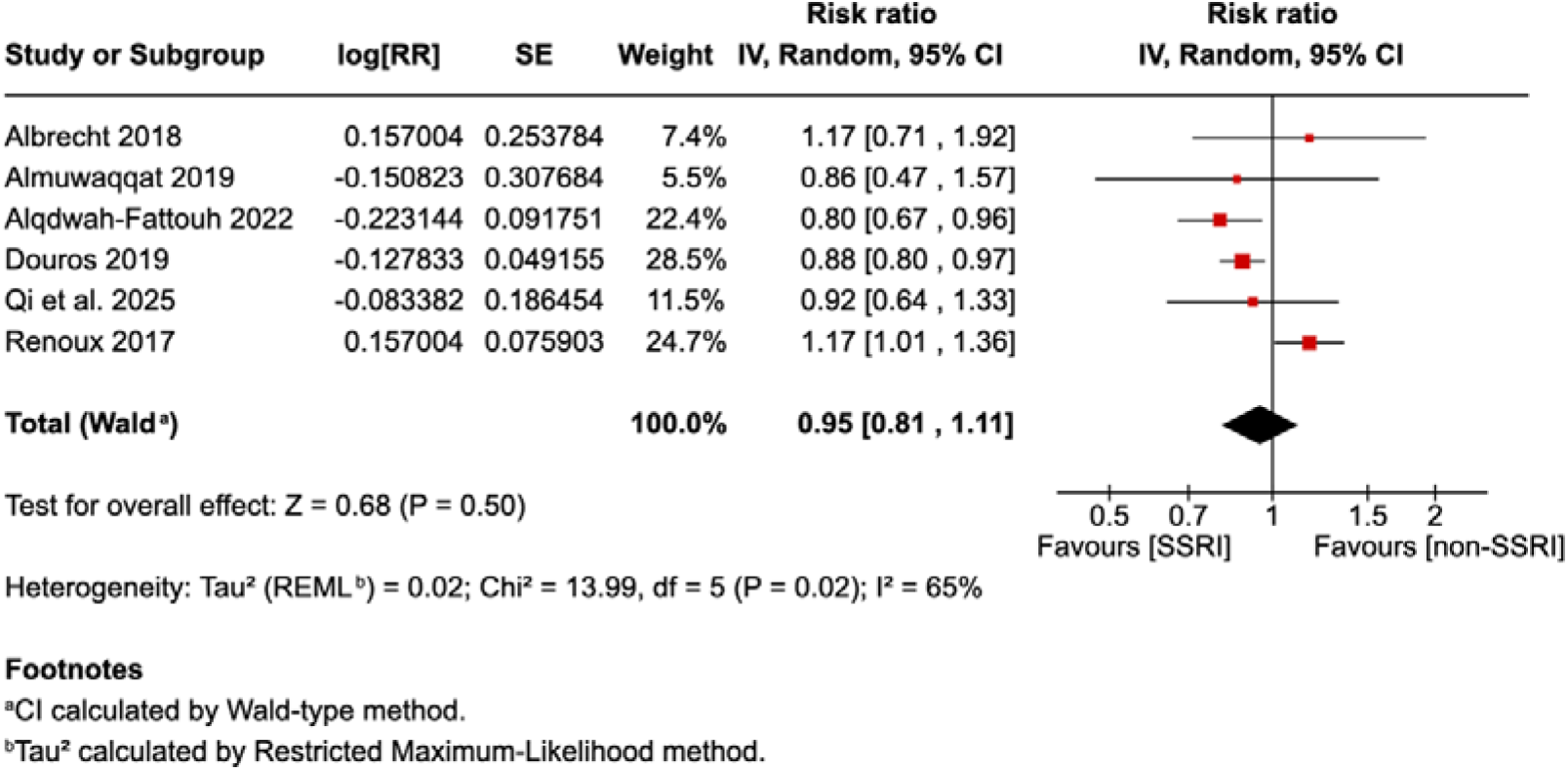
Forest plot showing RRs generated from retrieved observational studies (*n* = 5) and newly conducted retrospective cohort study. For Albrecht et al., the RR using SNRI as comparator and hemorrhagic stroke as outcome is used.

**Fig.**
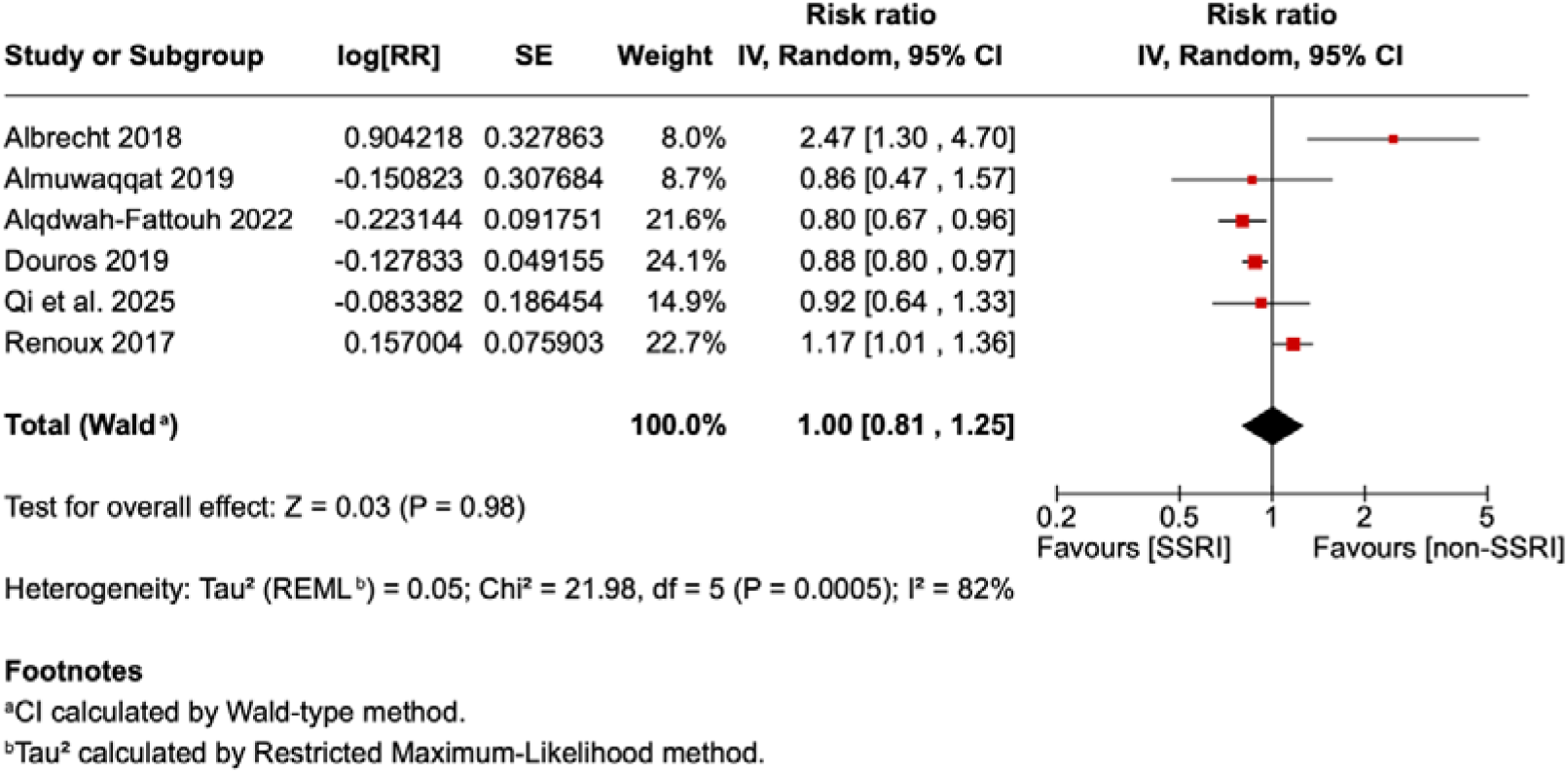
Forest plot showing RRs generated from retrieved observational studies (*n* = 5) and newly conducted retrospective cohort study. For Albrecht et al., the RR using TCA as comparator and hemorrhagic stroke as outcome is used.

**Fig.**
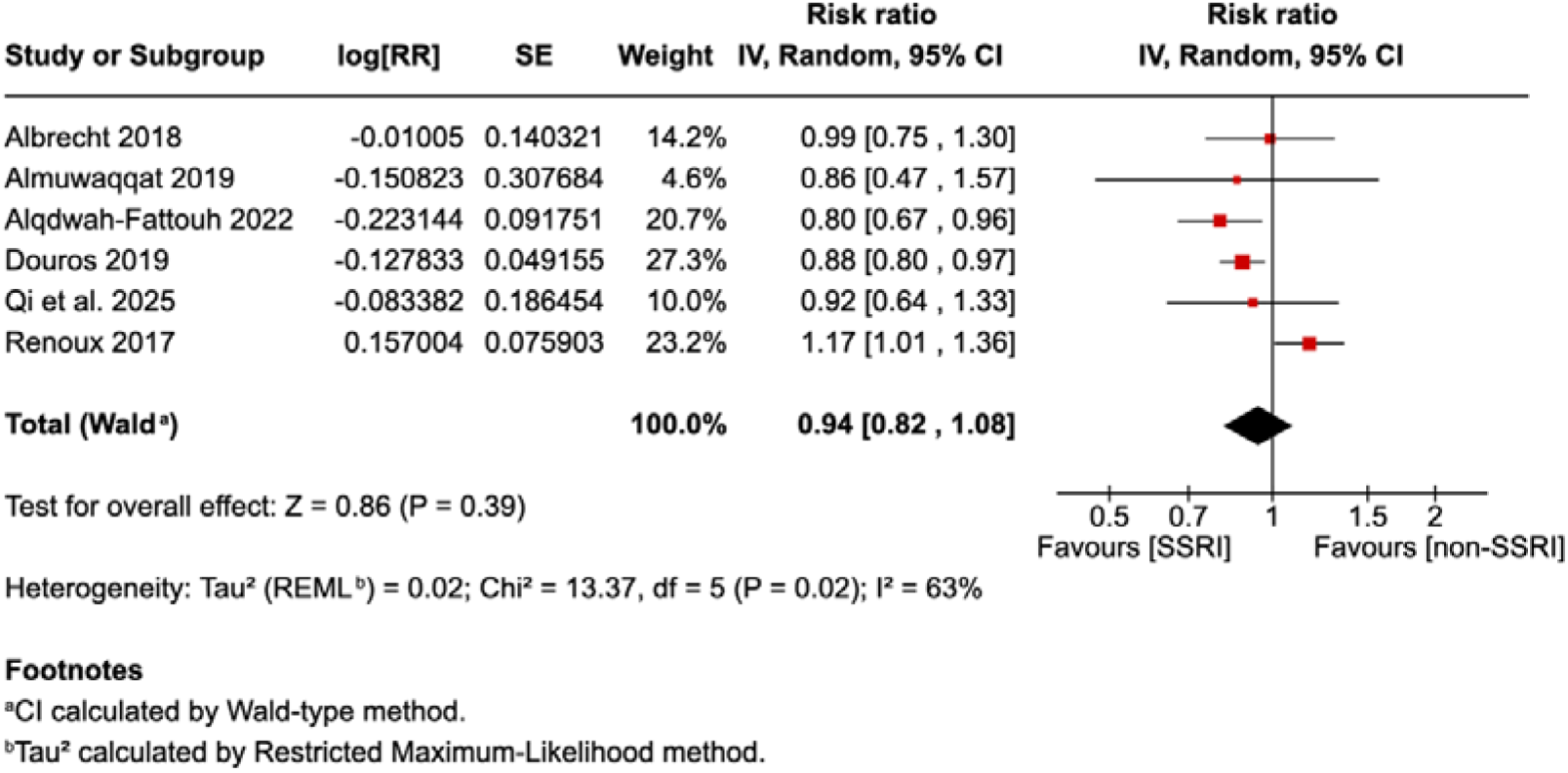
Forest plot showing RRs generated from retrieved observational studies (*n* = 5) and newly conducted retrospective cohort study. For Albrecht et al., the RR using TCA as comparator and ischemic stroke as outcome is used.

